# Estimating the Direct Impact of Influenza Vaccination and Vaccine-Averted Illnesses Using an Extended Static Compartmental Model

**DOI:** 10.64898/2026.10.23.26366241

**Authors:** Young M. Yoo, Anna N. Chard, Jorge H. Jara, Melissa A. Rolfes, Paula Couto, Francisco Nogareda, Lindsey M. Duca, Stefano Tempia

## Abstract

A previously developed static compartment model informed influenza program decisions by estimating the direct impact of influenza vaccination. To improve the estimation of the burden averted by vaccination, we extended this model to consider multiple influenza seasons, allow natural and vaccine-induced immunity to wane over time allow influenza vaccine effectiveness (VE) to differ by hospitalized versus non-hospitalized influenza-associated illness, and measure deaths averted. For model demonstration, we created a synthetic Northern Hemisphere-like and year-round influenza activity dataset spanning four seasons among older adults (>65 years) and assumed VE against symptomatic illness of 37% (17%–53%) and VE against hospitalization of 45% (28%–59%). Under Northern Hemisphere-like activity and an existing vaccination program with an average annual vaccine coverage of 33%, vaccination averted 15.1% (95% UI: 12.2%–16.3%) of influenza-associated illnesses across four seasons; the annual prevented fraction for hospitalized deaths ranged from 14.4% (95% UI: 11.7%–16.8%) to 18.5% (95% UI: 14.9%–21.5%). Under a hypothetical year round vaccination program with an annual coverage of 20%, vaccination averted 4.9% (95% UI: 3.8%–5.4%) of influenza-associated illnesses across four seasons; the annual prevented fraction for hospitalized deaths ranged from 5.5% (95% UI: 4.3%–6.5%) and 5.8% (95% UI: 4.8%–6.6%). These model extensions allow estimation of vaccine-averted burden across multiple seasons and diverse vaccination program settings, providing information to support influenza vaccination decision-making.

## INTRODUCTION

Influenza is a global public health threat, with seasonal epidemics resulting in substantial impacts on health, the economy, and society. An estimated 291,000 to 645,000 influenza-associated respiratory deaths occur globally each year [1]. Seasonal influenza vaccination is an effective method of preventing influenza-associated illnesses and its serious complications such as hospitalization and death [2,3]. Modeling studies can provide insights into the benefits of vaccination by quantifying the public health value of influenza vaccination programs. Such analyses have suggested that substantial influenza burden can be averted through vaccination in target populations, such as young children and pregnant women [4–6]. These estimates inform plausible and effective vaccination strategies, including timing of vaccination campaigns and deployment strategies, and for evaluating both existing and hypothetical vaccination programs across subpopulations of interest and time [7]. In turn, they provide public health officials with quantitative estimates to inform decisions about whether to introduce vaccination programs and how to implement to ensure timely protection as influenza activity increases. However, generating such estimates can be challenging in settings without access to specialized infectious disease modeling expertise. Approaches that can be implemented by national public health programs using locally available data may therefore be particularly valuable for translating surveillance data into evidence for vaccination policy and program decisions.

For more than a decade, the United States Centers for Disease Control and Prevention (U.S. CDC) has used a static compartmental model [7] to estimate the direct impact of annual influenza vaccination [8–11]. In 2018, Tokars, et al., evaluated this static compartmental model and further refined its implementation to omit an immune-lag adjustment to generate more accurate estimates of influenza cases averted by vaccination when working with monthly data [12]. To expand its use, in 2024 the U.S. CDC collaborated with the World Health Organization (WHO) and the Pan American Health Organization (PAHO) to develop a point-and-click Microsoft (MS) Excel tool based on the model [13], with a background chapter in the WHO Burden of Disease manual describing its methodology [14]. This simple, transparent, and user-oriented tool enables Ministry of Health officials and other public health practitioners to generate and interpret country-specific estimates of vaccination impact using their own surveillance and vaccination program data, without requiring specialized expertise in transmission modeling. This allows countries to directly evaluate and communicate the impact of their vaccination programs and generate evidence to inform decisions about program introduction, adjustment, or expansion. It has been used to estimate the burden of disease averted by influenza vaccination in several countries across the South American region [4,5].

Since then, many countries have improved their estimates for key parameters in the model (e.g., influenza-associated disease burden, vaccination coverage, vaccine effectiveness), highlighting growing demand for estimates of the burden averted by influenza vaccination. Previous versions of the static model were primarily designed for evaluating vaccination during a single influenza season in settings with seasonal, single-peak influenza circulation.

We addressed some of the limitations of this initial model in an updated framework that can be applied to a wider range of epidemiologic and programmatic settings by incorporating waning immunity [15,16], multi-season analyses, differential protection against severe disease [17], and mortality outcomes. In this paper, we present the extended model framework and illustrate its application under seasonal and year-round influenza activity patterns as well as existing and hypothetical influenza vaccination programs. We implemented these extensions while retaining the model’s relatively simple and transparent structure in a publicly available MS Excel tool, with the goal of enabling national public health programs to generate and explore vaccine-averted burden estimates using locally available data.

## METHODS

### Overview of model framework

Kostova, et al., and Tokars, et al. established a framework to estimate the direct impact of influenza vaccination on influenza-associated outcomes averted [7,12]. The model operated on a single-season basis; for multi-year analyses, it modeled each season separately. Under an all-or-none vaccination assumption, vaccine effectiveness (VE) against symptomatic illness is used to partition the vaccinated population into two distinct groups: effectively vaccinated and not effectively vaccinated.

These classifications refer to modeled protection against symptomatic illness, not to how the vaccine was administered. Effectively vaccinated individuals were conferred full protection immediately post-vaccination, but individuals that were not effectively vaccinated followed the same pathways as the non-vaccinated susceptible population and shared the same risk of infection. The model assumed that individuals were immune for the remainder of the season once infected or vaccinated, with no waning of immunity within the modeled period. We refer to this baseline model as the single-season, no-waning model throughout the paper.

We built our model extension upon this framework (**Figure 1**). The initial target population was assumed to be non-vaccinated and susceptible. Susceptible individuals who became infected experienced either non-hospitalized illness (i.e., symptomatic cases without hospitalization) or hospitalized illness, and a fraction of the influenza illnesses resulted in influenza-associated death. Non-hospitalized and hospitalized individuals returned to the non-vaccinated susceptible population as immunity waned. Each influenza-associated death was offset by the entry of one new susceptible individual to keep the target population size fixed. We considered this simplification reasonable because influenza-associated deaths were rare relative to the overall target population and were therefore unlikely to materially affect model estimates.

**Figure 1.**
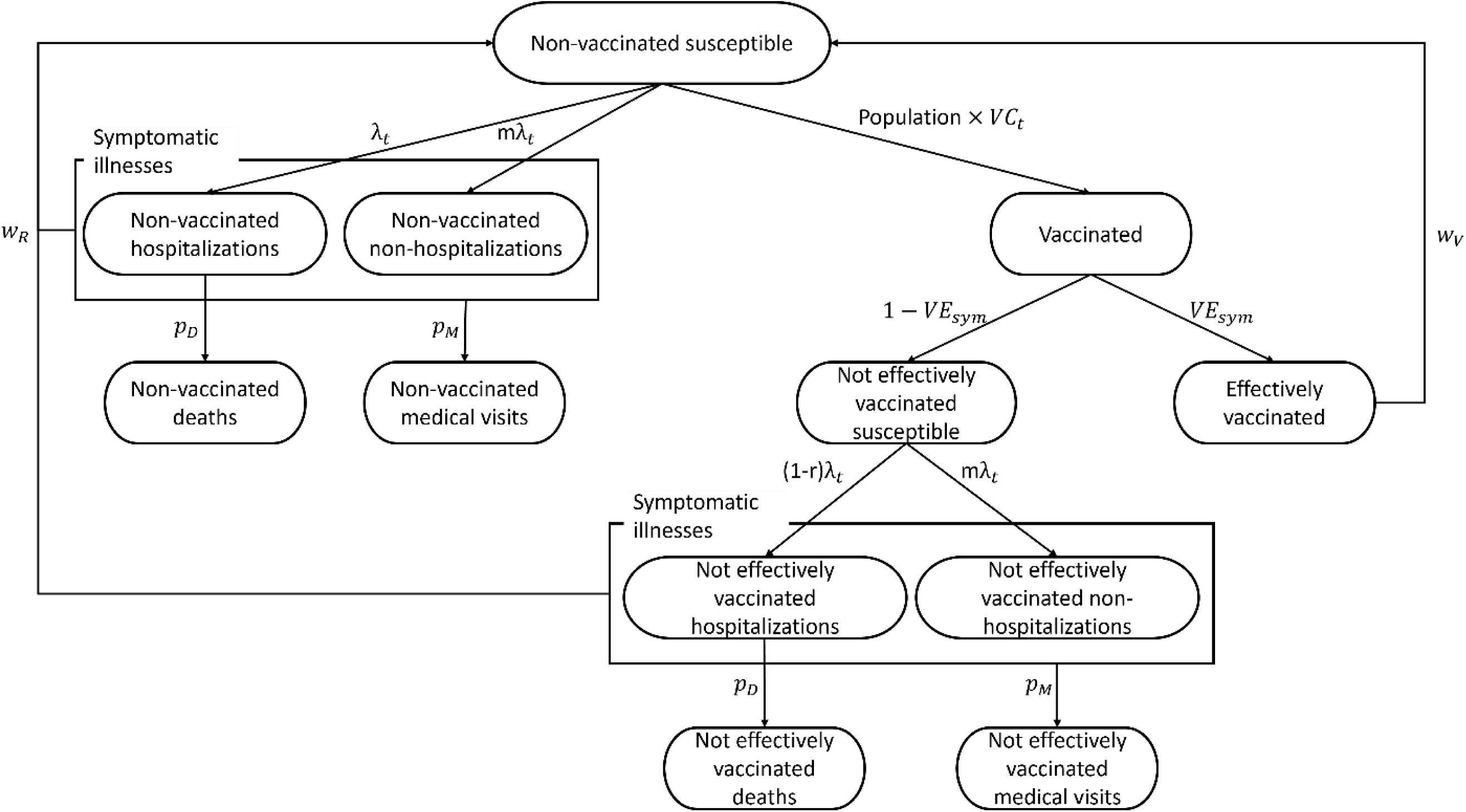
Structure and population pathways of the extended static compartmental model for estimating direct influenza vaccination impact in the target population. Parameter labels denote transition rates or proportions, with corresponding definitions provided in Table 1.

**Table 1.** Model input variables.

| Parameter | Parameter definition | Specification | Value used for model demonstration |
| --- | --- | --- | --- |
| $\lambda_t$ | Monthly hospitalization rate | $\frac{\text{Hospitalized Influenza Cases}_t}{\text{Susceptible Population}_{t-1}}$ | Monthly values provided in Supplementary Tables 1–2 |
| $m$ | Multiplier for ratio of hospitalized to non-hospitalized influenza-associated illnesses | Fixed input parameter, based on published estimates | 17.21 [1] |
| $p_M$ | Proportion of illnesses that were medically attended | Fixed input parameter, based on published estimates | 0.3969 [1] |
| $p_D$ | Proportion of influenza-associated hospitalizations resulting in death | Fixed input parameter, based on published estimates | 0.557 [2] |
| $w_R$ | Natural infection-induced immunity duration | Fixed input parameter set to 8 months | Applied across all modelled seasons |
| **Vaccination-related parameters** |  |  |  |
| $VC_t$ | Monthly vaccine coverage (%) | Time-varying input parameter, based on observed or hypothetical vaccine coverage data | Monthly values provided in Supplementary Tables 1–2 |
| $VE_{sym}$ | VE against symptomatic illness (%) | Fixed input parameter, based on published estimates | 37% (95% CI: 17% - 53%) [3] |
| $VE_{hosp}$ | VE against hospitalization (%) | Fixed input parameter, based on published estimates | 45% (95% CI: 28% - 59%) [3] |
| $r$ | Reduction in hospitalization risk (%) | $1 - \frac{(1 - VE_{hosp})}{(1 - VE_{sym})}$ | Derived using the VE values above |
| $w_V$ | Waning of vaccine-induced immunity | $\underline{VE}_t = VE - (1.37 \cdot 2 \cdot t) + (0.18 \cdot 2^2 \cdot t^2) - (0.03 \cdot 2^3 \cdot t^3)$ | Applied across all modelled seasons |

“t” indicates current month, “t-1” indicates prior month; VE, vaccine effectiveness; $\underline{VE}_t$ refers to vaccine effectiveness at month t, waning to 0% within approximately 8 months after vaccination

Each month, a proportion of the target population was vaccinated based on monthly vaccine coverage rates. We assumed that vaccination occurred at random, such that the likelihood of being vaccinated was independent of an individual’s exposure to, or susceptibility to, infection. This approach reflects how programs measure vaccine coverage in practice, where the denominator was the total population and ensured that the cumulative number of individuals vaccinated by the end of the vaccination campaign matched reported coverage. Among those vaccinated, effectively vaccinated individuals were assumed to be fully protected against symptomatic illness beginning in the month of vaccination. Effectively vaccinated individuals gradually returned to the non-vaccinated susceptible population as vaccine-induced immunity waned. Although influenza vaccination may not prevent symptomatic illness in all recipients, it may attenuate disease severity and reduce the likelihood of severe outcomes such as hospitalization [17,18]. To reflect this, individuals who were not effectively vaccinated shared the same risk of experiencing non-hospitalized illness as unvaccinated individuals but a lower risk of hospitalized illness. The model therefore included separate susceptible compartments for unvaccinated individuals and those who were not effectively vaccinated.

The model used target group-specific monthly influenza-associated hospitalization rates derived from routine national surveillance data as the primary input (**Table 1**). The non-hospitalized illness-to-hospitalization ratio and the proportion of hospitalizations resulting in death were applied to the monthly hospitalization rates to estimate non-hospitalized influenza illness and influenza-associated deaths, respectively. Medically attended illnesses were estimated by applying the proportion of influenza illnesses that were medically attended to the total monthly number of hospitalized and non-hospitalized illnesses. The model used two VE measures, which could be drawn from the literature or estimated in the target population: VE against symptomatic illness (*VE_sym_*), typically derived from outpatient studies, and VE against hospitalization (*VE*_*hosp*_), often based on data from severe acute respiratory infection surveillance. As in the single-season, no-waning model, *VE_sym_* was used to allocate the vaccinated population into effectively and not effectively vaccinated groups upon vaccination. Among individuals who were not effectively vaccinated against symptomatic illness, *VE_sym_* and *VE*_*hosp*_ were used to derive the reduction in hospitalization risk. Individuals who recovered from infection and those who were vaccinated returned to the susceptible population at rates *w_R_* and *w_V_*, respectively.

Our extension of the single-season, no-waning model allowed for season-specific input parameters to be applied to each 12-month interval. For instance, VE estimates may vary year-to-year due to differences in antigenic match between influenza vaccine-viruses and circulating influenza viruses in the community. Other input parameters, such as the hospitalization multiplier, may also vary by season.

### Expanded time horizon beyond 12-months with waning immunity

We extended the single-season, no-waning model beyond the 12-month time horizon of the baseline model; in the updated MS Excel tool, the model can be run for up to 48 months. Restricting the analysis to a single-season may fail to capture the full effects of vaccine protection, particularly when evaluating a year-round vaccine campaign in countries with year-round influenza circulation [19]. In these settings, infections and vaccinations that occurred later in the season could confer protection that is carried into the subsequent season. If we assumed that the entire target population was susceptible at the start of the analysis period, burden estimates in the first modeled year may be inflated. To address this issue, we introduced an initialization period (i.e., year-0), defined as the 12 months preceding the modeled period, which carried immunity forward into year 1. When year-0 specific inputs were unavailable, we approximated year-0 inputs using the seasons included in the analysis by default. We set hospitalizations in year 0 to the average across modeled seasons. We set year-0 vaccination coverage to 0% when the modeled period represented program initiation; when we modeled an existing vaccination program, we set year-0 coverage to the average coverage across the modeled seasons. We specified VE and other model inputs in year-0 using average values over the seasons included. When available, year-0 specific inputs were used in place of the default values.

Extending the time horizon beyond 12 months required specifying assumptions about the duration of immunity. Vaccine-induced immunity declined gradually with time elapsed since vaccination. As the protective effect of vaccination wanes, individuals who were initially effectively vaccinated gradually return to the susceptible population. Effectively vaccinated individuals were tracked by month of vaccination such that each monthly vaccination cohort followed its own waning trajectory based on time since vaccination. We parameterized waning VE as a function of time since vaccination using the cubic function proposed by Ferdinands, et al., which approximated exponential decay (**Equation 1**) [15]. Under this specification, VE declined to 0% within approximately 8 months after vaccination, with the exact timing dependent on the initial VE[19]. Any calculated VE values below 0% were set to 0%. For consistency with this waning profile and to adopt a conservative assumption [16], we assumed infection-acquired immunity persisted for the same 8-month period, after which individuals returned to the susceptible population.

**Equation 1**. **Estimating vaccine effectiveness t months after vaccination with waning vaccine-induced immunity**

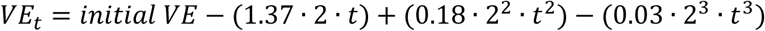

VE was typically assessed over the entire influenza season, which we assumed spanned 8 months, and could be thought of as an average VE for that season. Thus, if we modelled VE as declining from month to month, starting our calculations with the seasonal VE could lead to an underestimation of the vaccine’s overall effectiveness. To address this, we started with a higher initial VE which, after factoring in the decline over subsequent months, averaged out to match the reported seasonal VE. Initial VE was calibrated by minimizing the absolute difference between the mean of the cubic waning curve over the 8-month period and the reported VE. The tool also included an option to apply the reported seasonal VE as a constant value over the 8-month period rather than assuming any waning.

### Differential vaccine effectiveness for hospitalized versus non-hospitalized influenza illness

Influenza vaccination may offer varying levels of protection against different health outcomes. While receipt of the influenza vaccine may not completely prevent symptomatic infection, it may attenuate disease severity and reduce the likelihood of severe outcomes such as hospitalization [17,18]. To capture this potential benefit while retaining the structure of the original model framework, effectively vaccinated individuals were assumed to be fully protected against symptomatic illness, whereas individuals who were not effectively vaccinated remained at risk of symptomatic illness but could still have a reduced risk of hospitalization. Because *VE_hosp_* was typically estimated by comparing hospitalization risk in vaccinated versus unvaccinated individuals, it was not applied directly to the not effectively vaccinated group. Instead, we used *VE*_*hosp*_ and *VE_sym_* to derive the reduction in hospitalization risk among this group (**Equation 2**). This approach assumes that *VE*_*hosp*_ is at least as large as *VE_sym_* to allow for a reduced risk of hospitalization among individuals who were not effectively vaccinated. When *VE*_*hosp*_ < *VE_sym_*, we set the reduction in hospitalization risk to zero, equivalent to setting *VE*_*hosp*_ = *VE_sym_*. Thus, individuals who were not effectively vaccinated against symptomatic illness received no additional protection against hospitalization.

**Equation 2. Reduction in hospitalization risk for individuals not effectively vaccinated**

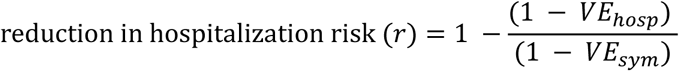

Individuals who were not effectively vaccinated were tracked by month of vaccination so that their reduction in hospitalization risk waned according to time since vaccination. For each monthly vaccination cohort, the reduction in hospitalization risk was recalculated in each subsequent month using the corresponding waned values of *VEsym* and *VE*_*hosp*_ (**Equation 3**). As vaccine-induced protection waned, this reduction approached zero, at which point individuals who were not effectively vaccinated had the same hospitalization risk as unvaccinated individuals.

**Equation 3: Estimating the reduction in hospitalization risk for individuals not effectively vaccinated *t* months after vaccination**

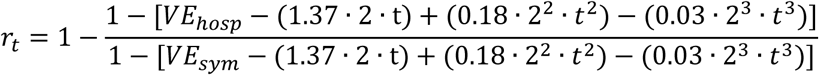

### Estimation of averted deaths

Depending on data availability, we estimated deaths averted by vaccination for one or both outcomes: 1) in-hospital influenza deaths, and 2) all influenza-associated deaths. In-hospital influenza deaths were estimated from observed influenza-associated hospitalizations using a multiplier approach [20]. Specifically, the number of in-hospital deaths was estimated by multiplying hospitalized influenza illnesses by the proportion of influenza-associated hospitalizations that resulted in death. This approach assumes that influenza-associated hospitalizations were appropriately estimated and that the proportion of hospitalizations resulting in death was applicable to the target population during the modeled period.

When data were available to account for influenza-associated deaths occurring outside the hospital setting, estimated in-hospital deaths were extrapolated to all influenza-associated deaths rather than modeled as a separate compartmental pathway. This extrapolation used national vital records data by multiplying the estimated number of in-hospital deaths by the inverse of the proportion of influenza-associated deaths that occurred in hospital among all influenza-coded deaths. This adjustment captured both individuals who were hospitalized at some point and those who were never hospitalized. This extrapolation assumes that the place-of-death multiplier was applicable to the modeled population and period that hospitalization surveillance and vital records data were sufficiently consistent.

### Overview of model outputs

For existing vaccination programs, the model estimated the counterfactual burden of influenza disease by severity (e.g., non-hospitalized illnesses, medically attended illnesses, hospitalized illnesses, and deaths) in the absence of vaccination. For this counterfactual scenario, vaccination coverage was set to 0%, such that the entire population followed the non-vaccinated pathways. The model then calculated averted burden as the difference between the hypothetical burden without vaccination and the observed burden with vaccination in place. For hypothetical vaccination programs, the model estimated the expected burden of influenza disease by severity under the hypothetical vaccination scenario and calculated averted burden as the difference between the observed burden without vaccination and the expected burden. We quantified uncertainty in model estimates using Monte Carlo simulation with 5,000 iterations. In each iteration, monthly hospitalizations were first summed to obtain annual hospitalized illnesses. Expected annual counts of non-hospitalized illnesses, medically attended illnesses, and deaths were then calculated by applying the corresponding multipliers to annual hospitalized illnesses. Annual counts for each outcome were sampled from Poisson distributions, and the outcome multipliers were recalculated from these sample counts and used in that iteration. Sampled annual hospitalized illnesses were then distributed across months according to the observed monthly distribution of hospitalizations. Similarly, the annual number of vaccinated individuals was sampled from a binomial distribution based on the target population size and annual vaccination coverage and then distributed across months according to the observed monthly distribution of vaccine coverage. VE estimates were sampled from a Beta-PERT distribution based on the point estimate and 95% confidence interval for each year or season. In addition to reporting averted burden as an absolute number, we calculated the prevented fraction, expressed as the proportion of averted outcomes to the potential outcomes in the absence of vaccination. We also reported the number needed to vaccinate (NNV) to avert an influenza-associated event, calculated as the total number vaccinated divided by the total number of cases averted. The previously developed MS Excel tool was updated to reflect the model extensions described in this paper and is available with supporting R code at https://github.com/CDCgov/PhaseII-Averted-Burden.

### Scenario analyses

As with the single-season, no waning model, the extended model framework maintained evaluation of alternative vaccination scenarios to assess how different deployment strategies influence the estimated averted burden under the same underlying disease burden. Users could specify alternative monthly vaccination coverage while holding other model inputs constant. For existing vaccination programs, the estimated monthly hospitalization rates in the absence of vaccination were used as the underlying disease burden, and alternative vaccination coverage scenarios were evaluated as counterfactual programs. For hypothetical vaccination programs, the observed burden was based on no vaccination, and alternative scenarios were evaluated by changing only the monthly vaccination coverage inputs.

### Synthetic dataset for model demonstration

To demonstrate use of the tool and illustrate model outputs, we generated two synthetic datasets, each covering a 48-month period. One dataset represented a setting with seasonal influenza activity typical in the Northern Hemisphere and was used to evaluate averted illness under an existing vaccination program with annual vaccine coverage of 28% to 37% distributed across October through January. The second dataset represented a setting with year-round influenza activity and no existing vaccination program. This dataset was used to evaluate averted illnesses that would occur under a hypothetical vaccination program with annual vaccine coverage of 20%, evenly distributed throughout the year. Throughout the modeled period, we assumed VE against symptomatic illness of 37% (17%–53%) and VE against hospitalization of 45% (28%–59%) [21]. The target age group for both datasets was older adults, and hospitalization rates were drawn from global average estimates ranging from 57 to 164 per 100,000 population, with values selected within this range [22]. The non-hospitalized illness-to-hospitalization ratio of 17.21 and the proportion of symptomatic illnesses that were medically attended (0.3969) were derived from U.S. influenza burden estimates over six seasons, from 2010–2011 season through 2015–2016 season [23]. The proportion of hospitalized illnesses resulting in death (0.0557) was derived from influenza hospitalizations and in-hospital deaths over three U.S. influenza seasons, from 2010–2011 through 2012–2013 [24]. We assumed a total target population of 600,000 individuals.

### Model validation

To assess model validity, we estimated vaccine-averted illnesses, medical attended illnesses, hospitalizations, and deaths using data from the 2017–2018 and 2018–2019 U.S. influenza seasons and compared our estimates with previously published age-specific estimates. Analyses were conducted separately for five age groups: 6 months–4 years, 5–17 years, 18–49 years, 50–64 years, and ≥65 years. Monthly influenza-associated hospitalization rates were obtained from FluSurv-NET [25], adjusted for under-detection, and extrapolated to the U.S. population within each age group. Outcome multipliers were obtained from the same published analyses used for comparison. Monthly age-specific vaccination coverage was obtained from FluVaxView [26]. We used age-specific VE estimates for each season, using overall influenza VE rather than virus-specific VE.

### Ethics

This activity was reviewed by the US Centers for Disease Control and Prevention (CDC), deemed to not constitute human subjects research, and conducted consistent with applicable federal law and CDC policy.

## RESULTS

We summarized the synthetic dataset inputs used for model demonstration (**Table 1 and Supplementary Tables 1–2**). Over the four modeled years, we based the synthetic Northern Hemisphere-like dataset on 2,868 influenza-associated hospitalizations, with activity starting around November and tapering off through March and April (**Figure 2A**). With existence of an influenza vaccine program, the estimated number of influenza-associated illnesses in the synthetic Northern Hemisphere-like dataset ranged from 9,898 (95% UI: 9,569–10,591) to 17,872 (95% UI: 17,335–19,086) across the four seasons (**Table 2**). Vaccination averted a total of 9,572 (95% UI: 7,666–10,422) influenza-associated illnesses with an average annual vaccine coverage of 33%. Across the four modeled seasons, the estimated seasonal averted burden ranged from 6–10 hospitalized deaths, 99–207 hospitalized illnesses, 623–1,302 medically-attended illnesses, and 1,471–3,073 non-hospitalized illnesses. Across seasons, 12.5% (95% UI: 8.7%–14.9%) to 16.1% (95% UI: 11.2%–19.3%) of all influenza illnesses estimated in the absence of vaccination were averted by vaccination. Estimates of the number needed to vaccinate to avert one influenza-associated illness varied by season, ranging from 62 (95% UI: 51–91) to 110 (95% UI: 90–159) persons.

**Figure 2.**
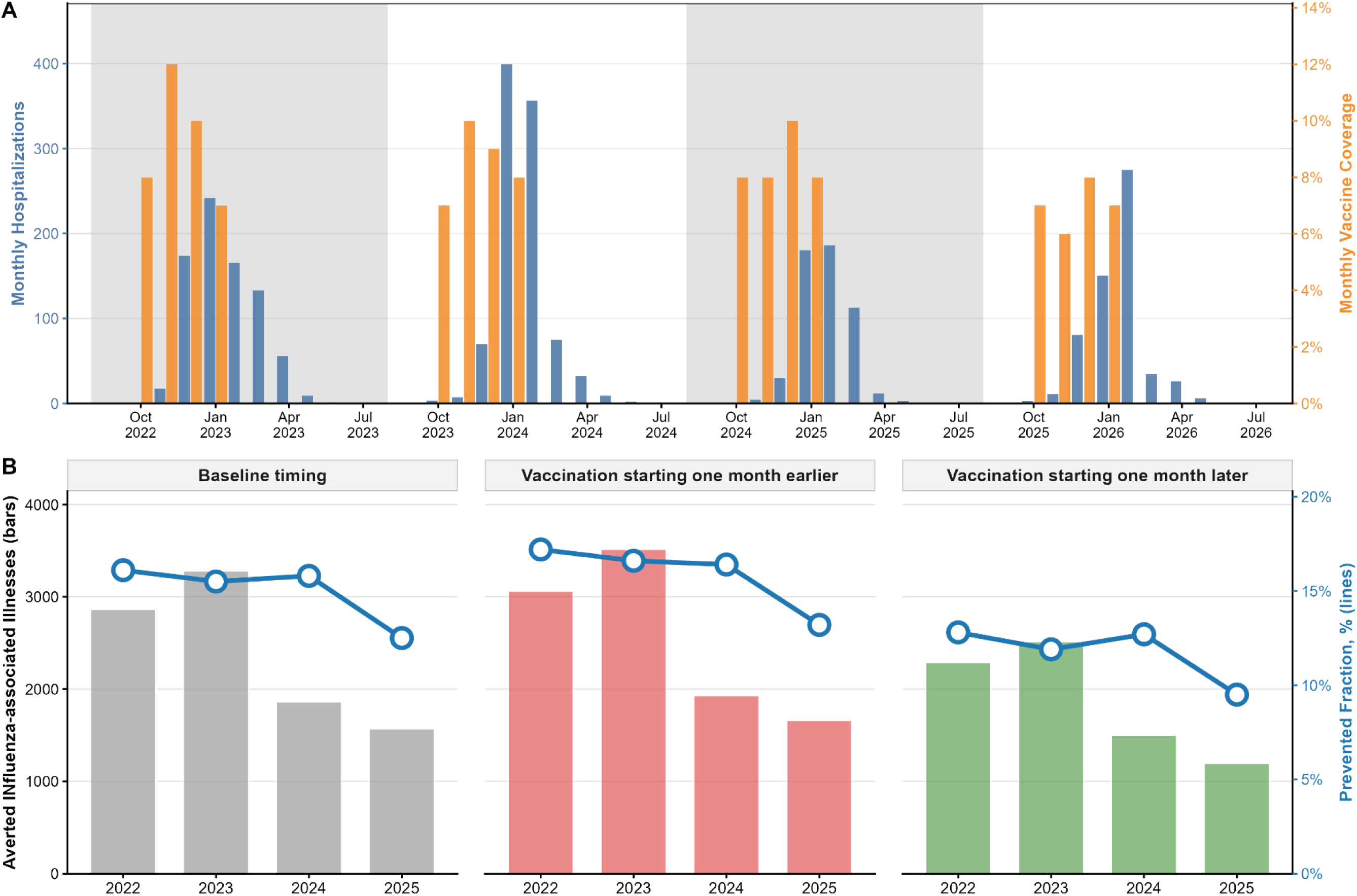
Synthetic Northern Hemisphere-like influenza activity, vaccine coverage, and estimated averted burden under alternative vaccination scenarios. A. Monthly hospitalizations and vaccine coverage used as model inputs. B. Annual influenza-associated illnesses averted as an absolute measure and prevented fraction as a relative measure of model-estimated impact.

**Table 2.** Estimated influenza-associated outcomes and averted burden under a current vaccination program in a Northern Hemisphere-like country.

| Season | Outcomes | Presence of vaccine (95% UI) | Absence of vaccine (95% UI) | Averted (95% UI) | Prevented fraction (95% UI) | NNV (95% UI) |
| --- | --- | --- | --- | --- | --- | --- |
| Season 1 | Non-hospitalized illnesses | 14,168 (13,707-15,197) | 16,850 (16,078-17,551) | 2,682 (1,801-3,308) | 15.9% (10.8%-19.2%) | 83 (67-123) |
|  | Medically-attended illnesses | 5,941 (5,724-6,368) | 7,077 (6,728-7,407) | 1,136 (781-1,402) | 16.1% (11.2%-19.3%) | 195 (158-284) |
|  | Hospitalized illnesses | 798 (745-854) | 979 (904-1,058) | 181 (139-221) | 18.5% (14.9%-21.5%) | 1,226 (1,005-1,599) |
|  | Hospitalized deaths | 44 (32-58) | 55 (39-71) | 10 (7-14) | 18.5% (14.9%-21.5%) | 22,001 (15,764-33,501) |
|  | Overall illnesses | 14,966 (14,497 - 15,991) | 17,829 (17,014 - 18,568) | 2,863 (1,968 -3,515) | 16.1% (11.2%-19.3%) | 78 (63-113) |
| Season 2 | Non-hospitalized illnesses | 16,918 (16,391-18,133) | 19,991 (19,133-20,773) | 3,073 (2,042-3,754) | 15.4% (10.4%-18.4%) | 66 (54-100) |
|  | Medically-attended illnesses | 7,094 (6,847-7,605) | 8,396 (8,007-8,766) | 1,302 (888-1,577) | 15.5% (10.7%-18.4%) | 157 (129-230) |
|  | Hospitalized illnesses | 954 (893-1,015) | 1,161 (1,075-1,247) | 207 (160-251) | 17.9% (14.5%-20.7%) | 984 (813-1,275) |
|  | Hospitalized deaths | 53 (39-68) | 65 (48-83) | 12 (8-16) | 17.9% (14.5%-20.7%) | 17,650 (12,896-26,009) |
|  | Overall illnesses | 17,872 (17,335 - 19,086) | 21,152 (20,255 - 21,977) | 3,280 (2,233 -3,978) | 15.5% (10.7%-18.4%) | 62 (51-91) |
| Season 3 | Non-hospitalized illnesses | 9,370 (9,047-10,063) | 11,112 (10,602-11,588) | 1,742 (1,172-2,127) | 15.7% (10.7%-18.8%) | 117 (96-174) |
|  | Medically-attended illnesses | 3,929 (3,770-4,222) | 4,667 (4,436-4,899) | 738 (510-898) | 15.8% (11.0%-18.8%) | 276 (227-400) |
|  | Hospitalized illnesses | 528 (483-573) | 646 (584-707) | 118 (89-144) | 18.2% (14.7%-21.1%) | 1,736 (1,416-2,285) |
|  | Hospitalized deaths | 29 (19-40) | 36 (23-50) | 7 (4-9) | 18.2% (14.7%-21.1%) | 31,142 (21,561-51,448) |
|  | Overall illnesses | 9,898 (9,569 -10,591) | 11,757 (11,217 - 12,261) | 1,859 (1,279 -2,258) | 15.8% (11.0%-18.8%) | 110 (90-159) |
| Season 4 | Non-hospitalized illnesses | 10,359 (10,067-10,942) | 11,830 (11,402-12,230) | 1,471 (985-1,783) | 12.4% (8.4%-14.9%) | 114 (94-170) |
|  | Medically-attended illnesses | 4,345<br>(4,191-4,609) | 4,969<br>(4,759-5,182) | 623<br>(427-754) | 12.5%<br>(8.7%-14.9%) | 270<br>(223-393) |
|  | Hospitalized illnesses | 588<br>(542-635) | 687<br>(630-748) | 99<br>(76-120) | 14.4%<br>(11.7%-16.8%) | 1,692<br>(1,397-2,198) |
|  | Hospitalized deaths | 33<br>(22-44) | 38<br>(26-52) | 6<br>(3-8) | 14.4%<br>(11.7%-16.8%) | 30,360<br>(21,157-48,082) |
|  | Overall illnesses | 10,947<br>(10,646 - 11,537) | 12,518<br>(12,073 - 12,945) | 1,570<br>(1,073-1,896) | 12.5%<br>(8.7%-14.9%) | 107<br>(89-156) |
| All seasons | Non-hospitalized illnesses | 50,816<br>(50,090-52,885) | 59,783<br>(58,448-61,080) | 8,967<br>(7,091-9,780) | 15.0%<br>(12.0%-16.2%) | 89<br>(82-113) |
|  | Medically-attended illnesses | 21,309<br>(20,971-22,185) | 25,109<br>(24,518-25,717) | 3,800<br>(3,047-4,138) | 15.1%<br>(12.2%-16.3%) | 210<br>(193-262) |
|  | Hospitalized illnesses | 2,868<br>(2,762-2,971) | 3,473<br>(3,330-3,620) | 605<br>(530-679) | 17.4%<br>(15.7%-19.0%) | 1,318<br>(1,174-1,507) |
|  | Hospitalized deaths | 160<br>(136-185) | 194<br>(164-225) | 34<br>(27-41) | 17.4%<br>(15.7%-19.0%) | 23,656<br>(19,649-29,036) |
|  | Overall illnesses | 53,684<br>(52,940 - 55,766) | 63,256<br>(61,838 - 64,611) | 9,572<br>(7,666 - 10,422) | 15.1%<br>(12.2%-16.3%) | 83<br>(77-104) |

Abbreviation: NNV, number needed to vaccinate; UI, uncertainty interval.

The synthetic year-round influenza activity dataset included 2,580 influenza-associated hospitalizations in the absence of vaccination (**Figure 3A**). During four years, the estimated number of influenza-associated illnesses in the synthetic dataset with year-round influenza activity ranged from 6,885 (95% UI: 6,720–7,044) to 17,157 (95% UI: 16,902–17,408) per year in the absence of vaccination (**Table 3**). Under a hypothetical year-round vaccination program with an annual coverage of 20%, a total of 2,311 (95% UI: 1,788–2,530) influenza-associated illnesses were averted over four years. The estimated yearly averted burden ranged from 1–3 hospitalized deaths, 22–55 hospitalized illnesses, 136– 339 medically-attended illnesses, and 321–800 non-hospitalized illnesses. Although the number of illnesses averted varied year-to-year, vaccination averted a similar proportion of influenza illnesses estimated in the absence of vaccination across years, between 4.8% (95% UI: 3.1%–5.9%) and 5% (95% UI: 3.4%–5.8%). Estimates of the number needed to vaccinate to avert one influenza-associated illness varied by year, ranging from 140 persons (95% UI: 121–201) to 350 persons (95% UI: 305–493). One of the key features of this tool is that it allows users to evaluate alternative vaccine campaign deployment strategies under the same underlying disease burden by varying the timing and monthly distribution of vaccine coverage. For example, using the synthetic Northern Hemisphere-like dataset, we evaluated vaccination beginning one month earlier or one month later than the baseline strategy (**Figure 2B**). Starting the vaccine campaign one month earlier resulted in 6.2% additional influenza-associated illnesses averted across all seasons. Using the synthetic year-round influenza activity dataset, we compared year-round vaccine deployment with seasonal campaigns timed to periods typical Southern and Northern Hemisphere vaccination program (**Figure 3B–3C**). The seasonal campaign starting in April resulted in 22.6% additional influenza-associated illnesses averted across all years. These examples illustrate how the tool can be used to quantify the impact of changes in vaccine delivery relative to influenza activity.

**Figure 3.**
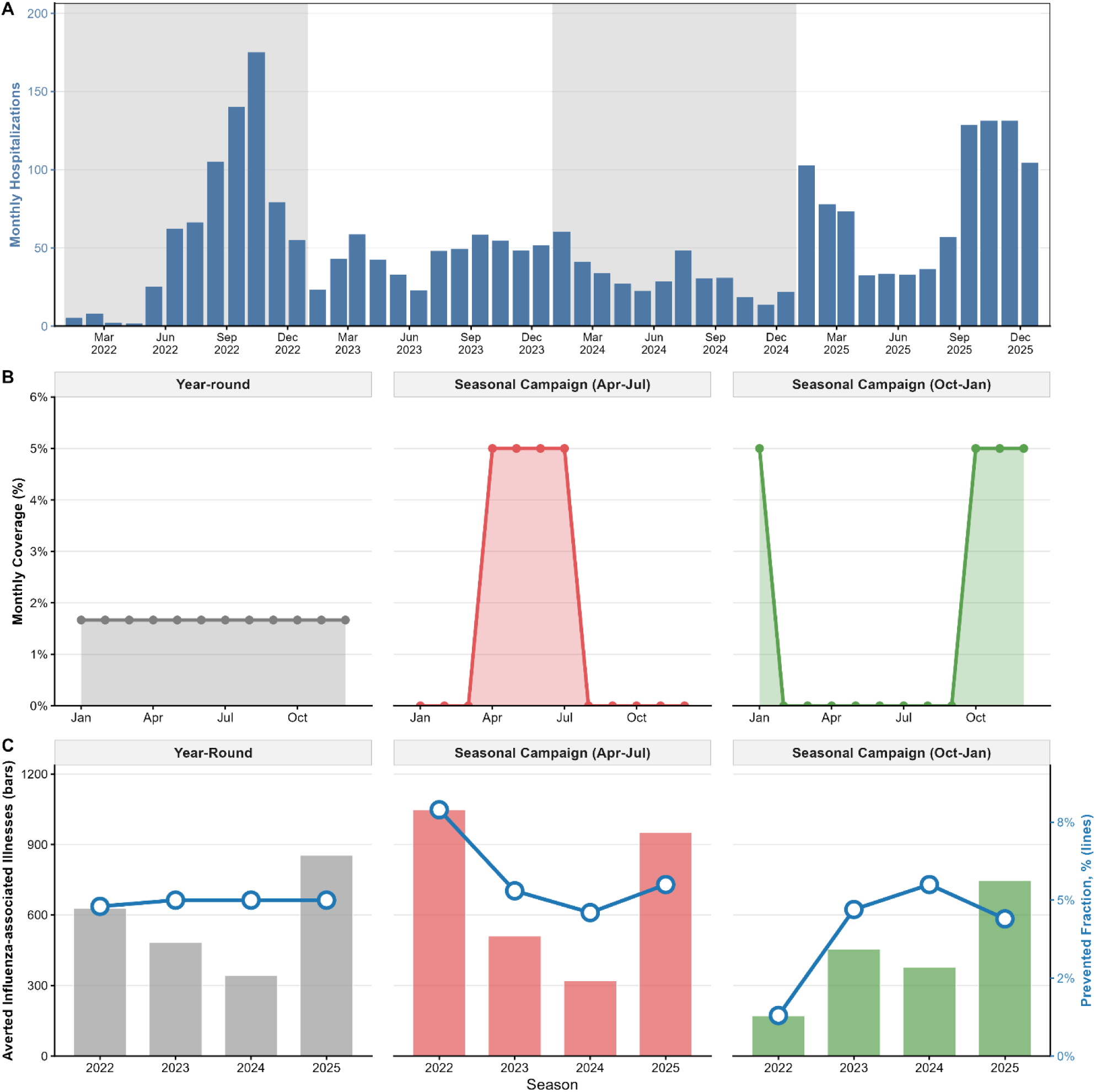
Synthetic year-round influenza activity, vaccine coverage, and estimated averted burden under alternative vaccination scenarios. A. Monthly hospitalizations in the absence of a vaccine program. B. Vaccination deployment strategies for a hypothetical annual vaccine coverage of 20%. C. Annual influenza-associated illnesses averted as an absolute measure and prevented fraction as a relative measure of model-estimated impact.

**Table 3.**
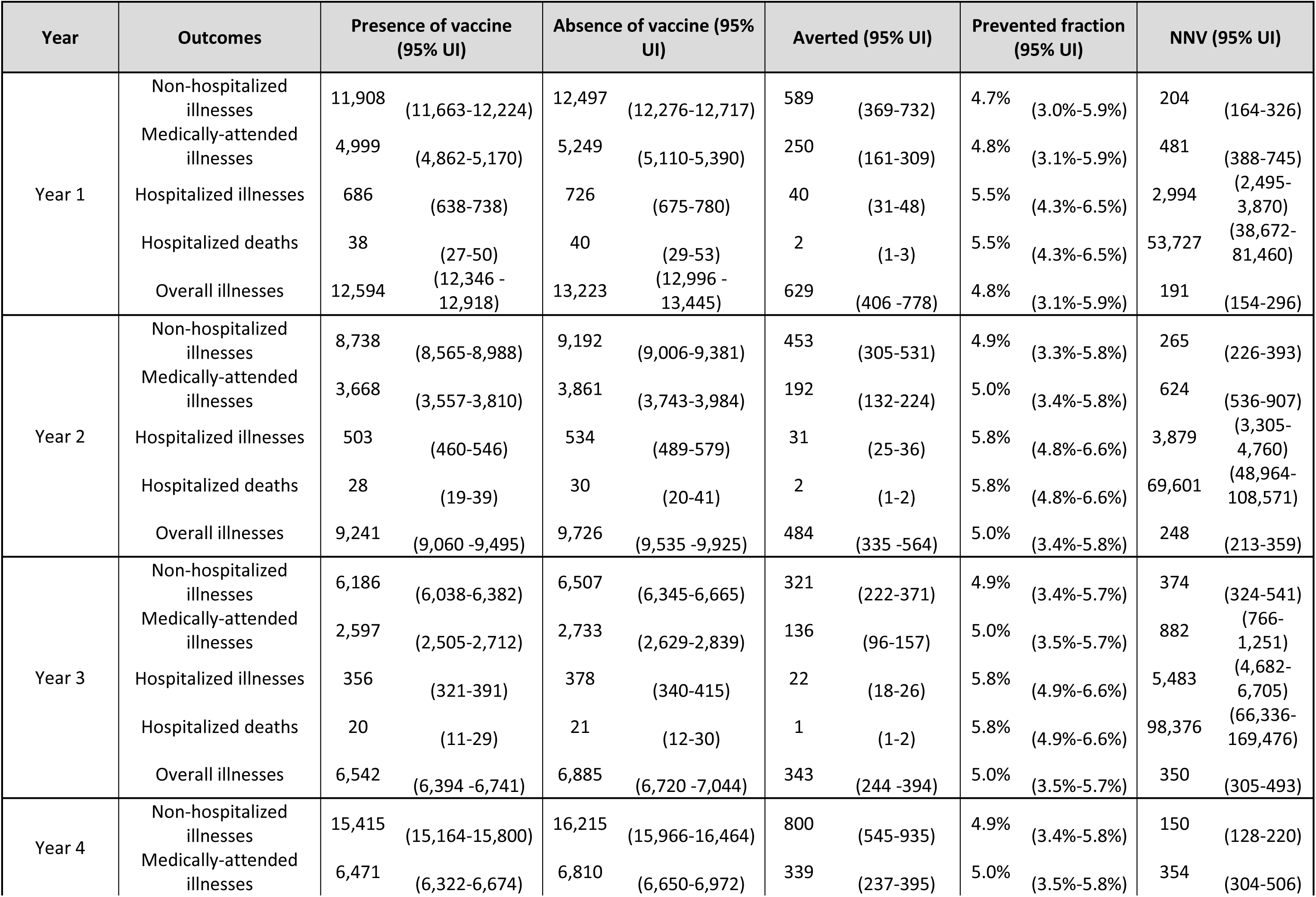
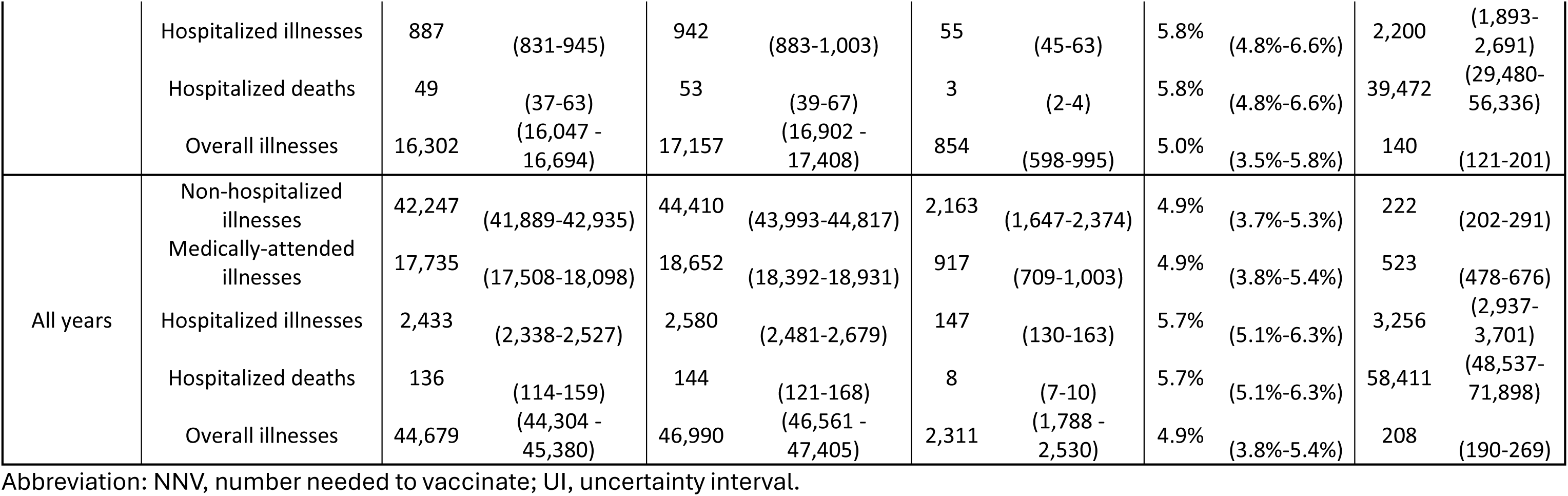
Estimated influenza-associated outcomes and averted burden under a hypothetical year-round vaccination program in a country with year-round influenza transmission.

As part of model validation, averted burden estimates from the extended model were broadly comparable in magnitude to the previously published estimates (**Figure 4 and Supplementary Tables 3– 6**). In the 2017–2018 season, point estimates under the waning VE assumption were consistently higher than the published estimates across age groups and outcomes, although their 95% uncertainty intervals overlapped. Estimates under the constant VE assumption were generally closer to the published estimates. In the 2018–2019 season, point estimates under both VE assumptions were within the 95% uncertainty intervals of the published estimates across age groups and outcomes. For the 6 months–4 years, 50–64 years, and ≥65 years age groups, the 95% uncertainty intervals under both VE assumptions were within the corresponding published uncertainty intervals.

**Figure 4.**
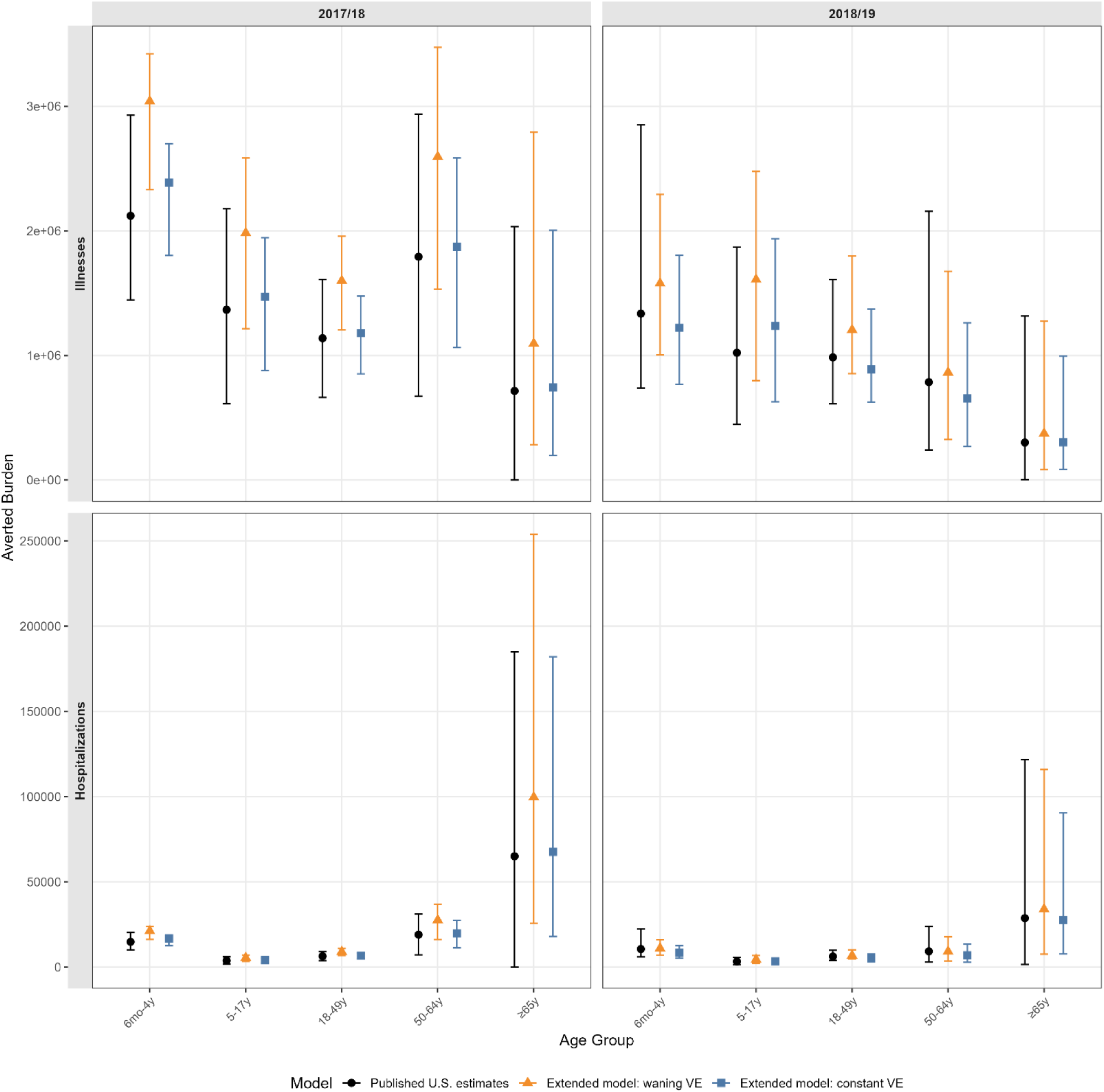
Comparison of averted burden estimates from the extended model and previously published U.S. estimates, 2017–2018 and 2018–2019 influenza seasons.

## DISCUSSION

A previously developed static compartment model has been helpful in informing influenza program decisions related to vaccine introduction, expansion, and timing. In this paper, we extended the model in several ways. Collectively, these extensions allow users to evaluate vaccination programs in settings where influenza circulation occurs throughout the year or over multiple years, reflect real-world understanding of waning immunity over time, and variable performance of vaccine by disease severity. We also added the ability to estimate the number of influenza-associated deaths averted, aligning model outputs with commonly reported burden estimates [27]. These adaptations, along with the ability to explore both existing and hypothetical alternate vaccine strategies, allow the model to better reflect how influenza circulates and how vaccination programs operate in real-world settings.

Importantly, these additional capabilities were incorporated while retaining a relatively simple and transparent model structure. The model is intended to provide a practical approach for national public health programs to generate estimates of the direct impact of vaccination using locally available data, without requiring specialized expertise in transmission modeling. Implementation in an accessible MS Excel tool allows users to examine model inputs and assumptions, modify vaccination program scenarios, and generate locally relevant estimates. Additionally, expressing results in terms of prevented fraction and the number needed to vaccinate complements estimates of averted burden and supports communication of findings, particularly since absolute averted burden is sensitive to the underlying influenza attack rate [7]. This combination of expanded analytic capabilities and accessibility enables countries to generate and interpret their own estimates to support vaccination program decisions.

The previous Excel tool implementing the single-season, no-waning model has been used to estimate averted illness in low- and middle-income countries [4,5]. In parallel, countries have expanded their capacity to generate influenza burden estimates. Strengthened sentinel respiratory disease surveillance systems and standardized methods described in the WHO Burden of Disease manual have supported generation of national influenza burden estimates [28,29], while PAHO’s Network for the Evaluation of Vaccine Effectiveness in Latin America and the Caribbean-influenza (la Red para la Evaluación de Vacunas en Latino América y el Caribe—influenza [REVELAC–i]) consistently generates and publishes influenza vaccine effectiveness estimates for key target populations, providing critical inputs for the model [30–32]. By incorporating the model extensions described in this paper, national programs can bring these locally generated data together with an updated tool that better reflects changes in immunity and protection over time, addresses local program needs, and supports routine evaluation of influenza vaccine programs. In response to growing interest in such evaluations, PAHO hosted a workshop in Brazil in 2026 that trained 36 representatives from 13 countries on the use of the extended averted burden MS Excel tool, strengthening local analytic capacity [33].

In model validation to published U.S. estimates, estimates derived from the expanded model were consistently within the 95% uncertainty intervals of the published estimates, giving us confidence in the validity of this tool. The higher averted burden estimates under the waning VE assumption likely reflect, in part, the calibration of the waning curve, which results in an initial VE higher than the reported seasonal VE. During these seasons, most vaccination occurred before the main period of seasonal influenza activity, consistent with a typical Northern Hemisphere vaccination campaign. As a result, periods of higher VE soon after vaccination overlapped with influenza circulation, resulting in greater estimated protection than under the constant VE assumption. Under the constant VE assumption, estimates were close to those from the U.S. estimates but narrower uncertainty intervals. This reflected differences in how uncertainty was sampled and in how influenza was modeled.

Our model is subject to several limitations. First, model estimates may be sensitive to input values because hospitalization rates from surveillance data were scaled to estimate target population burden. For example, influenza mortality burden derived from this hospitalization-based multiplier approach may underestimate the true burden if the multipliers do not fully capture under-ascertainment of influenza-associated deaths [34]. The more closely the multipliers reflect the health care access, care-seeking behavior, surveillance practices, and other societal and contextual differences, the more accurate the estimates are likely to be. Despite this limitation, averted burden estimates may still provide useful approximation of direct vaccination impact.

Second, this model was designed as an extension of the single-season, no-waning model framework. As a result, we did not include a transmission rate, nor did we account for incubation, infectious, and recovery periods that reflect the natural progression of the disease, as is typical in dynamic compartmental models. Instead, the model used a monthly time step and incidence rates, providing a snapshot of the disease burden in each month. This represents an intentional tradeoff between model complexity and accessibility. Dynamic transmission models are better suited to questions requiring estimation of transmission dynamics, indirect effects, or interactions among population groups. In contrast, the static framework requires fewer assumptions and specialized inputs and can be implemented and interpreted by public health programs using routinely available data.

Third, we did not account for the indirect effects of vaccination, such as potential protection of unvaccinated individuals through herd immunity. Although indirect effects can contribute to the overall impact of influenza vaccination, their magnitude remains uncertain because data quantifying indirect protection are limited and may vary by transmission intensity and characteristics of the population [35]. The averted burden, prevented fraction, and number needed to vaccinate produced by the model reflect only the direct impact of vaccination. Nevertheless, direct impact is an important and readily interpretable measure of vaccination program benefit and can provide decision makers with locally relevant estimates of illnesses, hospitalizations, and deaths prevented through vaccination. As the estimates captured direct effects of influenza vaccination and the impact of vaccination is non-linear [36], the results represent minimum estimates of vaccine impact.

Fourth, influenza vaccination may attenuate illness severity even when infection occurred [17]. However, a previous meta-analysis found that VE estimates from inpatient settings were not consistently higher than those from outpatient settings [37]. A U.S. study comparing VE in inpatient and outpatient networks reported higher inpatient VE in one season, but this pattern was not consistent across years [38]. Differences in clinical case definitions, enrollment criteria, and underlying patient characteristics may result in different patient populations across settings and could have contributed to these findings. When available evidence indicates that the VE against hospitalization is lower than that against symptomatic illness, we assumed no additional reduction in hospitalization risk among individuals who were not effectively vaccinated.

Fifth, natural immunity was modeled as providing protection against influenza for eight months, after which individuals returned to the susceptible population without progressive waning. This assumption did not allow reinfection during the protected period and the model treated infection-derived immunity the same regardless of influenza types or subtypes. Some cohort studies suggested that reinfections with influenza viruses were possible within the same season [39–41]. Because circulating influenza types and subtypes can differ both within and between seasons, this simplification may overstate infection-derived protection. Also, post-infection seroprotection may persist for much of a season or longer [16]. If prior infection provided protection for longer than assumed in the model, estimates of vaccine-averted burden could be overestimated. Lastly, the model assumed a waning rate for vaccine-induced immunity derived from prior analyses of VE among older adults in the U.S [15]. Prior work suggested a broadly similar pattern of waning VE among children and older adults [42]. We also assumed that VE against severe and non-severe illnesses waned at the same rate, although these may differ in actuality. In addition, vaccine-induced protection that had not yet waned was carried across seasons without accounting for antigenic drift.

## CONCLUSION

This work expands a previously developed model for estimating vaccine-averted influenza burden by allowing users to evaluate influenza vaccination programs across multiple seasons, account for waning immunity, consider differences in protection against influenza-associated severe and non-severe outcomes, and estimate deaths averted. By better reflecting key features of influenza epidemiology and vaccination programs while retaining a transparent and accessible model structure, the extended model provides national public health programs with a practical approach to generate their own estimates of vaccine-averted burden using locally available data and to evaluate and communicate the impact of influenza vaccination strategies.

## Data Availability

The model described in this manuscript is implemented in a publicly available tool at

https://github.com/CDCgov/PhaseII-Averted-Burden

## ACKNOWLEDGEMENT

We wish to thank Chiedza Machingaidze for conceptualization. We wish to thank Karen R. Border and Matthew Biggerstaff for critical subject matter review.

## DISCLAIMER

The findings and conclusions in this report are those of the authors and do not necessarily represent the official position of the Centers for Disease Control and Prevention.

## REFERENCE

1. Iuliano AD, Roguski KM, Chang HH, Muscatello DJ, Palekar R, Tempia S, et al. Estimates of global seasonal influenza-associated respiratory mortality: a modelling study. The Lancet. 2018 Mar 31;391(10127):1285–300. doi:10.1016/S0140-6736(17)33293-2

2. Minozzi S, Lytras T, Gianola S, Gonzalez-Lorenzo M, Castellini G, Galli C, et al. Comparative efficacy and safety of vaccines to prevent seasonal influenza: A systematic review and network meta-analysis. eClinicalMedicine. 2022 Apr 1;46. doi:10.1016/j.eclinm.2022.101331 PubMed PMID: 35360146.

3. CDC. Influenza (Flu) [Internet]. 2026 [cited 2026 Sep 22]. Key Facts About Seasonal Flu Vaccine. Available from: https://www.cdc.gov/flu/vaccines/keyfacts.html

4. Chard AN, Machingaidze C, Loayza S, Gharpure R, Nogareda F, González R, et al. Estimating averted illnesses from influenza vaccination for children and pregnant women – El Salvador, Panama, and Peru, 2011–2018. Vaccine. 2024 Apr 6. doi:10.1016/j.vaccine.2024.04.007

5. Jara JH, Loayza S, Nogareda F, Couto P, Descalzo MA, Chard AN, et al. Influenza Vaccine-Averted Illness in Chile, Guyana, and Paraguay During 2013–2018: A Standardized Approach to Assess the Value of Vaccination. J Infect Dis. 2025 Mar 15;231(Supplement_2):S133–43. doi:10.1093/infdis/jiaf038

6. Morris SE, O’Halloran A, Sundaresan D, Olson SM, Chung JR, Ellington S, et al. Public health benefits of maternal influenza vaccination among pregnant women and infants <6 months in the United States, 2011–2020. Vaccine. 2026 Aug 30;89:128895. doi:10.1016/j.vaccine.2026.128895

7. Kostova D, Reed C, Finelli L, Cheng PY, Gargiullo PM, Shay DK, et al. Influenza Illness and Hospitalizations Averted by Influenza Vaccination in the United States, 2005–2011. Goldstein E, editor. PLoS ONE. 2013 Jun 19;8(6):e66312. doi:10.1371/journal.pone.0066312

8. CDC. Flu Burden [Internet]. 2025 [cited 2026 Jan 27]. Flu Burden Prevented by Vaccination: Past Seasons. Available from: https://www.cdc.gov/flu-burden/php/data-vis-vac/past-burden-prevented-est.html

9. Rolfes MA, Flannery B, Chung JR, O’Halloran A, Garg S, Belongia EA, et al. Effects of Influenza Vaccination in the United States During the 2017–2018 Influenza Season. Clin Infect Dis. 2019 Nov 13;69(11):1845–53. doi:10.1093/cid/ciz075

10. Chung JR, Rolfes MA, Flannery B, Prasad P, O’Halloran A, Garg S, et al. Effects of Influenza Vaccination in the United States During the 2018–2019 Influenza Season. Clin Infect Dis. 2020 Nov 5;71(8):e368–76. doi:10.1093/cid/ciz1244

11. Morris SE, Mathis SM, Chung JR, Flannery B, O’Halloran A, Cummings CN, et al. Estimating historical disease burden and the impact of vaccination by influenza type and subtype in the United States, 2016–2020. Vaccine X. 2025 Oct 1;26:100700. doi:10.1016/j.jvacx.2025.100700

12. Tokars JI, Rolfes MA, Foppa IM, Reed C. An evaluation and update of methods for estimating the number of influenza cases averted by vaccination in the United States. Vaccine. 2018 Nov 19;36(48):7331–7. doi:10.1016/j.vaccine.2018.10.026 PubMed PMID: 30327213; PubMed Central PMCID: PMC6666399.

13. World Health Organization. ChiedzaWHO. Influenza-Burden-Burden-Averted [Internet]. 2024 [cited 2025 Aug 28]. Available from: https://github.com/ChiedzaWHO/Influenza-Burden-Burden-Averted

14. World Health Organization. An operational tool to estimate influenza disease burden averted through vaccination [Internet]. World Health Organization; 2025 [cited 2025 Sep 19]. Available from: https://iris.who.int/handle/10665/381767 doi:10.2471/B09385

15. Ferdinands JM, Alyanak E, Reed C, Fry AM. Waning of Influenza Vaccine Protection: Exploring the Trade-offs of Changes in Vaccination Timing Among Older Adults. Clin Infect Dis. 2020 Apr 10;70(8):1550–9. doi:10.1093/cid/ciz452

16. Hoa LNM, Sullivan SG, Mai LQ, Khvorov A, Phuong HVM, Hang NLK, et al. Influenza A(H1N1)pdm09 But Not A(H3N2) Virus Infection Induces Durable Seroprotection: Results From the Ha Nam Cohort. J Infect Dis. 2022 Jul 1;226(1):59–69. doi:10.1093/infdis/jiaa293

17. Regan AK, Arriola CS, Couto P, Duca L, Loayza S, Nogareda F, et al. Severity of influenza illness by seasonal influenza vaccination status among hospitalised patients in four South American countries, 2013–19: a surveillance-based cohort study. Lancet Infect Dis. 2023 Feb 1;23(2):222–32. doi:10.1016/S1473-3099(22)00493-5 PubMed PMID: 36206790.

18. Ferdinands JM, Thompson MG, Blanton L, Spencer S, Grant L, Fry AM. Does influenza vaccination attenuate the severity of breakthrough infections? A narrative review and recommendations for further research. Vaccine. 2021 Jun 23;39(28):3678–95. doi:10.1016/j.vaccine.2021.05.011

19. Gharpure R, Yoo YM, Andagalu B, Tempia S, Loayza S, Machingaidze C, et al. Estimating Influenza Illnesses Averted by year-Round and Seasonal Campaign Vaccination for Young Children, Kenya - volume 30, Number 11—November 2024 - Emerging Infectious Diseases journal - CDC [Internet]. doi:10.3201/eid3011.240375

20. Steele MK, Couture A, Reed C, Iuliano D, Whitaker M, Fast H, et al. Estimated Number of COVID-19 Infections, Hospitalizations, and Deaths Prevented Among Vaccinated Persons in the US, December 2020 to September 2021. JAMA Netw Open. 2022 Jul 6;5(7):e2220385. doi:10.1001/jamanetworkopen.2022.20385

21. Rose AM, Lucaccioni H, Marsh K, Kirsebom F, Whitaker H, Emborg HD, et al. Interim 2024/25 influenza vaccine effectiveness: eight European studies, September 2024 to January 2025. Eurosurveillance. 2025 Feb 20;30(7):2500102. doi:10.2807/1560-7917.ES.2025.30.7.2500102

22. Paget J, Staadegaard L, Wang X, Li Y, van Pomeren T, van Summeren J, et al. Global and national influenza-associated hospitalisation rates: Estimates for 40 countries and administrative regions. J Glob Health. 2023 Jan 27;13:04003. doi:10.7189/jogh.13.04003 PubMed PMID: 36701368; PubMed Central PMCID: PMC9879557.

23. Rolfes MA, Foppa IM, Garg S, Flannery B, Brammer L, Singleton JA, et al. Annual estimates of the burden of seasonal influenza in the United States: A tool for strengthening influenza surveillance and preparedness. Influenza Other Respir Viruses. 2018 Jan;12(1):132–7. doi:10.1111/irv.12486 PubMed PMID: 29446233; PubMed Central PMCID: PMC5818346.

24. Reed C, Chaves SS, Kirley PD, Emerson R, Aragon D, Hancock EB, et al. Estimating Influenza Disease Burden from Population-Based Surveillance Data in the United States. PLOS ONE. 2015 Mar 4;10(3):e0118369. doi:10.1371/journal.pone.0118369

25. CDC. FluView [Internet]. 2026 [cited 2026 Sep 24]. Influenza Hospitalization Surveillance Network (FluSurv-NET). Available from: https://www.cdc.gov/fluview/overview/influenza-hospitalization-surveillance.html

26. CDC. FluVaxView [Internet]. 2026 [cited 2026 Sep 22]. Influenza Vaccination Coverage for Persons 6 months and Older. Available from: https://www.cdc.gov/fluvaxview/interactive/general-population-coverage.html

27. Home | Seasonal Influenza Disease Burden Estimator [Internet]. [cited 2025 Sep 30]. Available from: https://www.flutool.org/

28. Lee VJ, Ho ZJM, Goh EH, Campbell H, Cohen C, Cozza V, et al. Advances in measuring influenza burden of disease. Influenza Other Respir Viruses. 2018;12(1):3–9. doi:10.1111/irv.12533

29. World Health Organization. A manual for estimating disease burden associated with seasonal influenza [Internet]. Geneva: World Health Organization; 2015 [cited 2024 Aug 10]. 124 p. Available from: https://iris.who.int/handle/10665/178801

30. Fowlkes AL. Interim Effectiveness Estimates of 2023 Southern Hemisphere Influenza Vaccines in Preventing Influenza-Associated Hospitalizations — REVELAC–i Network, March–July 2023. MMWR Morb Mortal Wkly Rep. 2023;72. doi:10.15585/mmwr.mm7237e1

31. Zeno EE. Interim Effectiveness Estimates of 2024 Southern Hemisphere Influenza Vaccines in Preventing Influenza-Associated Hospitalization — REVELAC-i Network, Five South American Countries, March–July 2024. MMWR Morb Mortal Wkly Rep. 2024;73. doi:10.15585/mmwr.mm7339a1

32. Russ S. Interim Effectiveness Estimates of 2025 Southern Hemisphere Influenza Vaccines in Preventing Influenza-Associated Outpatient and Hospitalized Illness — Eight Southern Hemisphere Countries, March–September 2025. MMWR Morb Mortal Wkly Rep. 2025;74. doi:10.15585/mmwr.mm7436a3

33. 33. Pan American Health Organization. Regional Workshop in Brazil Enhances Capacity to Measure Influenza Burden and Vaccination Impact - PAHO/WHO | Pan American Health Organization [Internet]. 2026 Feb 9 [cited 2026 Feb 18]. Available from: https://www.paho.org/en/news/9-2-2026-regional-workshop-brazil-enhances-capacity-measure-influenza-burden-and-vaccination

34. Li L, Wong JY, Wu P, Bond HS, Lau EHY, Sullivan SG, et al. Heterogeneity in Estimates of the Impact of Influenza on Population Mortality: A Systematic Review. Am J Epidemiol. 2018 Feb 1;187(2):378–88. doi:10.1093/aje/kwx270

35. Krauland MG, Mandell A, Roberts MS. Estimated Burden of Influenza and Direct and Indirect Benefits of Influenza Vaccination. JAMA Netw Open. 2025 Jul 16;8(7):e2521324. doi:10.1001/jamanetworkopen.2025.21324

36. Tuite AR, Fisman DN. Number-needed-to-vaccinate calculations: Fallacies associated with exclusion of transmission. Vaccine. 2013 Jan 30;31(6):973–8. doi:10.1016/j.vaccine.2012.11.097

37. Feng S, Cowling BJ, Sullivan SG. Influenza vaccine effectiveness by test-negative design – Comparison of inpatient and outpatient settings. Vaccine. 2016 Mar 29;34(14):1672–9. doi:10.1016/j.vaccine.2016.02.039

38. Tenforde MW, Chung J, Smith ER, Talbot HK, Trabue CH, Zimmerman RK, et al. Influenza Vaccine Effectiveness in Inpatient and Outpatient Settings in the United States, 2015–2018. Clin Infect Dis. 2021 Aug 1;73(3):386–92. doi:10.1093/cid/ciaa407

39. Zhang W, Gao J, Chen L, Tian J, Biggerstaff M, Zhou S, et al. Estimated influenza illnesses and hospitalizations averted by influenza vaccination among children aged 6–59 months in Suzhou, China, 2011/12 to 2015/16 influenza seasons. Vaccine. 2020 Dec 3;38(51):8200–5. doi:10.1016/j.vaccine.2020.10.069

40. Cohen C, Kleynhans J, Moyes J, McMorrow ML, Treurnicht FK, Hellferscee O, et al. Asymptomatic transmission and high community burden of seasonal influenza in an urban and a rural community in South Africa, 2017–18 (PHIRST): a population cohort study. Lancet Glob Health. 2021 Jun 1;9(6):e863–74. doi:10.1016/S2214-109X(21)00141-8

41. Horby P, Mai LQ, Fox A, Thai PQ, Thi Thu Yen N, Thanh LT, et al. The Epidemiology of Interpandemic and Pandemic Influenza in Vietnam, 2007–2010. Am J Epidemiol. 2012 May 15;175(10):1062–74. doi:10.1093/aje/kws121

42. Belongia EA, Sundaram ME, McClure DL, Meece JK, Ferdinands J, VanWormer JJ. Waning vaccine protection against influenza A (H3N2) illness in children and older adults during a single season. Vaccine. 2015 Jan 1;33(1):246–51. doi:10.1016/j.vaccine.2014.06.052

