## Supplementary Material for "Estimating the Direct Impact of Influenza Vaccination and Vaccine-Averted Illnesses Using an Extended Static Compartmental Model"

**SUPPLEMENTAL MATERIALS**

*Specification of synthetic dataset input values*

For the Northern Hemisphere seasonal dataset, monthly influenza-associated hospitalizations were distributed according to the proportional distribution of cases observed in FluNet, the influenza surveillance data reported to the WHO Global Influenza Surveillance and Response System, using data from the European region [1]. For the year-round influenza activity dataset, hospitalizations were generated using the same approach, but distributions were informed by data from Kenya, Thailand, Singapore and Viet Nam. Annual vaccination coverage estimates for older adults were obtained from the WHO/UNICEF Joint Reporting Form on Immunization [2].

Vaccine effectiveness inputs were obtained from interim European estimates, with VE against symptomatic illness from primary care studies and VE against hospitalization from hospital-based studies [3]. The non-hospitalized-to-hospitalized multiplier and the proportion of infections that were medically attended were drawn from a published study among older adults in the United States [4]. The proportion of hospitalizations resulting in influenza-associated death was similarly drawn from published U.S. estimates [5]. The same input values were used across modeled seasons and were applied to both synthetic datasets.

**Supplementary Table 1. Monthly input values for older adults (≥65 years) over the 48-month modeled period under Northern Hemisphere-like influenza activity**

|  | **Year 1** | | **Year 2** | | **Year 3** | | **Year 4** | |
| --- | --- | --- | --- | --- | --- | --- | --- | --- |
|  | **Monthly vaccine coverage** | **Monthly number of hospitalizations** | **Monthly vaccine coverage** | **Monthly number of hospitalizations** | **Monthly vaccine coverage** | **Monthly number of hospitalizations** | **Monthly vaccine coverage** | **Monthly number of hospitalizations** |
| August | 0% | 0 | 0% | 0 | 0% | 0 | 0% | 0 |
| September | 0% | 0 | 0% | 0 | 0% | 0 | 0% | 0 |
| October | 8% | 0 | 7% | 3 | 8% | 1 | 7% | 3 |
| November | 12% | 17 | 10% | 7 | 8% | 4 | 6% | 11 |
| December | 10% | 174 | 9% | 69 | 10% | 30 | 8% | 81 |
| January | 7% | 242 | 8% | 399 | 8% | 180 | 7% | 150 |
| February | 0% | 166 | 0% | 356 | 0% | 186 | 0% | 275 |
| March | 0% | 133 | 0% | 75 | 0% | 112 | 0% | 35 |
| April | 0% | 56 | 0% | 32 | 0% | 11 | 0% | 26 |
| May | 0% | 9 | 0% | 9 | 0% | 3 | 0% | 6 |
| June | 0% | 1 | 0% | 2 | 0% | 0 | 0% | 1 |
| July | 0% | 0 | 0% | 0 | 0% | 0 | 0% | 0 |

**Supplementary Table 2. Monthly input values for older adults (≥65 years) over the 48-month modeled period under year-round influenza activity**

|  | **Year 1** | | **Year 2** | | **Year 3** | | **Year 4** | |
| --- | --- | --- | --- | --- | --- | --- | --- | --- |
|  | **Monthly vaccine coverage** | **Monthly number of hospitalizations** | **Monthly vaccine coverage** | **Monthly number of hospitalizations** | **Monthly vaccine coverage** | **Monthly number of hospitalizations** | **Monthly vaccine coverage** | **Monthly number of hospitalizations** |
| January | 2% | 5 | 2% | 23 | 2% | 60 | 2% | 103 |
| February | 2% | 8 | 2% | 43 | 2% | 41 | 2% | 78 |
| March | 2% | 2 | 2% | 59 | 2% | 34 | 2% | 73 |
| April | 2% | 2 | 2% | 42 | 2% | 27 | 2% | 32 |
| May | 2% | 25 | 2% | 33 | 2% | 23 | 2% | 33 |
| June | 2% | 62 | 2% | 23 | 2% | 29 | 2% | 33 |
| July | 2% | 66 | 2% | 48 | 2% | 49 | 2% | 37 |
| August | 2% | 105 | 2% | 49 | 2% | 31 | 2% | 57 |
| September | 2% | 140 | 2% | 58 | 2% | 31 | 2% | 129 |
| October | 2% | 175 | 2% | 55 | 2% | 18 | 2% | 131 |
| November | 2% | 79 | 2% | 48 | 2% | 14 | 2% | 131 |
| December | 2% | 55 | 2% | 52 | 2% | 22 | 2% | 104 |

**Supplementary Table 3. Published U.S. and extended model estimates of averted influenza-associated illnesses with 95% uncertainty intervals under waning and constant VE assumptions, by influenza season and age group**

|  |  | **Published U.S. estimates** | | **Extended model: waning VE** | | **Extended model: constant VE** | |
| --- | --- | --- | --- | --- | --- | --- | --- |
|  |  | **Number prevented** | **95% UI** | **Number prevented** | **95% UI** | **Number prevented** | **95% UI** |
| 2017/18 | 6mo-4y | 2,121,511 | (1,445,133–2,928,929) | 3,041,563 | (2,331,045–3,420,706) | 2,388,927 | (1,802,834–2,698,943) |
|  | 5-17y | 1,366,965 | (613,310–2,178,412) | 1,983,495 | (1,214,461–2,587,271) | 1,471,687 | (880,274–1,944,770) |
|  | 18-49y | 1,138,407 | (663,181–1,610,481) | 1,600,296 | (1,206,981–1,957,274) | 1,180,608 | (852,234–1,476,634) |
|  | 50-64y | 1,792,530 | (673,687–2,937,768) | 2,595,267 | (1,531,479–3,473,692) | 1,873,158 | (1,063,404–2,585,949) |
|  | ≥65y | 715,073 | (0–2,033,756) | 1,096,274 | (282,669–2,792,933) | 743,646 | (198,138–2,004,666) |
| 2018/19 | 6mo-4y | 1,335,840 | (736,978–2,851,485) | 1,579,011 | (1,004,939–2,293,671) | 1,222,686 | (768,079–1,804,265) |
|  | 5-17y | 1,021,821 | (446,661–1,870,063) | 1,609,358 | (798,171–2,477,611) | 1,237,065 | (628,110–1,937,131) |
|  | 18-49y | 984,698 | (612,071–1,610,237) | 1,205,251 | (853,090–1,798,653) | 888,172 | (625,765–1,371,239) |
|  | 50-64y | 785,710 | (241,045–2,158,024) | 864,063 | (325,357–1,676,135) | 654,773 | (269,257–1,262,157) |
|  | ≥65y | 300,879 | (3,249–1,316,421) | 373,755 | (83,950–1,275,822) | 303,731 | (85,092–994,759) |

**Supplementary Table 4. Published U.S. and extended model estimates of averted medically attended illnesses with 95% uncertainty intervals under waning and constant VE assumptions, by influenza season and age group**

|  |  | **Published U.S. estimates** | | **Extended model: waning VE** | | **Extended model: constant VE** | |
| --- | --- | --- | --- | --- | --- | --- | --- |
|  |  | **Number prevented** | **95% UI** | **Number prevented** | **95% UI** | **Number prevented** | **95% UI** |
| 2017/18 | 6mo-4y | 1,421,413 | (971,080–1,966,976) | 2,037,847 | (1,562,003–2,292,253) | 1,600,581 | (1,208,254–1,808,460) |
|  | 5-17y | 710,822 | (319,168–1,143,256) | 1,031,417 | (631,741–1,345,700) | 765,277 | (457,650–1,011,519) |
|  | 18-49y | 421,211 | (243,149–603,887) | 592,109 | (446,605–724,434) | 436,825 | (315,380–546,714) |
|  | 50-64y | 770,788 | (292,197–1,263,230) | 1,115,965 | (658,451–1,493,605) | 805,458 | (457,486–1,111,847) |
|  | ≥65y | 400,441 | (0–1,145,616) | 613,914 | (158,375–1,564,246) | 416,442 | (110,942–1,123,008) |
| 2018/19 | 6mo-4y | 895,012 | (492,938–1,926,307) | 1,057,938 | (673,245–1,538,038) | 819,199 | (514,563–1,209,438) |
|  | 5-17y | 531,347 | (230,928–980,184) | 836,866 | (415,338–1,288,448) | 643,274 | (326,661–1,006,835) |
|  | 18-49y | 364,338 | (225,540–600,040) | 445,943 | (315,784–665,134) | 328,624 | (231,559–507,367) |
|  | 50-64y | 337,855 | (102,597–926,454) | 371,547 | (139,928–720,757) | 281,553 | (115,794–542,936) |
|  | ≥65y | 168,492 | (1,844–737,768) | 209,303 | (47,038–714,187) | 170,090 | (47,677–557,238) |

**Supplementary Table 5. Published U.S. and extended model estimates of averted hospitalizations with 95% uncertainty intervals under waning and constant VE assumptions, by influenza season and age group**

|  |  | **Published U.S. estimates** | | **Extended model: waning VE** | | **Extended model: constant VE** | |
| --- | --- | --- | --- | --- | --- | --- | --- |
|  |  | **Number prevented** | **95% UI** | **Number prevented** | **95% UI** | **Number prevented** | **95% UI** |
| 2017/18 | 6mo-4y | 14,790 | (10,075–20,419) | 21,210 | (16,265–23,865) | 16,659 | (12,584–18,821) |
|  | 5-17y | 3,748 | (1,682–5,973) | 5,439 | (3,324–7,084) | 4,035 | (2,412–5,325) |
|  | 18-49y | 6,390 | (3,722–9,040) | 8,980 | (6,773–10,999) | 6,625 | (4,785–8,308) |
|  | 50-64y | 19,009 | (7,144–31,154) | 27,521 | (16,216–36,856) | 19,864 | (11,279–27,403) |
|  | ≥65y | 65,007 | (0–184,887) | 99,661 | (25,717–253,804) | 67,604 | (18,004–182,067) |
| 2018/19 | 6mo-4y | 10,569 | (5,933–22,379) | 11,011 | (6,996–16,017) | 8,526 | (5,345–12,597) |
|  | 5-17y | 3,269 | (1,478–5,709) | 4,413 | (2,188–6,798) | 3,392 | (1,722–5,314) |
|  | 18-49y | 6,239 | (3,919–9,884) | 6,763 | (4,794–10,107) | 4,984 | (3,516–7,710) |
|  | 50-64y | 9,250 | (2,970–23,951) | 9,163 | (3,452–17,779) | 6,944 | (2,856–13,387) |
|  | ≥65y | 28,695 | (1,533–121,752) | 33,978 | (7,628–115,914) | 27,612 | (7,731–90,466) |

**Supplementary Table 6. Published U.S. and extended model estimates of averted deaths with 95% uncertainty intervals under waning and constant VE assumptions, by influenza season and age group**

|  |  | **Published U.S. estimates** | | **Extended model: waning VE** | | **Extended model: constant VE** | |
| --- | --- | --- | --- | --- | --- | --- | --- |
|  |  | **Number prevented** | **95% UI** | **Number prevented** | **95% UI** | **Number prevented** | **95% UI** |
| 2017/18 | 6mo-4y | 74 | (0–189) | 117 | (83–39) | 92 | (64–110) |
|  | 5-17y | 89 | (28–197) | 44 | (26–59) | 32 | (19–44) |
|  | 18-49y | 228 | (119–403) | 332 | (251–407) | 245 | (177–307) |
|  | 50-64y | 868 | (330–1,591) | 1,018 | (599–1,371) | 735 | (417–1,020) |
|  | ≥65y | 6,796 | (0–19,844) | 8,671 | (2,226–22,104) | 5,882 | (1,567–15,830) |
| 2018/19 | 6mo-4y | 111 | (25–381) | 116 | (72–171) | 90 | (55–134) |
|  | 5-17y | 33 | (4–108) | 44 | (22–70) | 34 | (17–54) |
|  | 18-49y | 229 | (114–588) | 248 | (175–370) | 182 | (128–282) |
|  | 50-64y | 536 | (143–2,000) | 531 | (201–1,025) | 402 | (166–771) |
|  | ≥65y | 2,625 | (122–12,163) | 3,109 | (697–10,651) | 2,526 | (706–8,317) |

**REFERENCE**

1.	World Health Organization. Flunet [Internet]. [cited 2026 Jan 29]. Available from: https://www.who.int/tools/flunet

2.	World Health Organization. Immunization Data [Internet]. [cited 2026 Jan 29]. WHO Immunization Data portal - All Data. Available from: https://immunizationdata.who.int/global

3.	Rose AM, Lucaccioni H, Marsh K, Kirsebom F, Whitaker H, Emborg HD, et al. Interim 2024/25 influenza vaccine effectiveness: eight European studies, September 2024 to January 2025. Eurosurveillance. 2025 Feb 20;30(7):2500102. doi:10.2807/1560-7917.ES.2025.30.7.2500102

4.	Rolfes MA, Foppa IM, Garg S, Flannery B, Brammer L, Singleton JA, et al. Annual estimates of the burden of seasonal influenza in the United States: A tool for strengthening influenza surveillance and preparedness. Influenza Other Respir Viruses. 2018 Jan;12(1):132–7. doi:10.1111/irv.12486 PubMed PMID: 29446233; PubMed Central PMCID: PMC5818346.

5.	Reed C, Chaves SS, Kirley PD, Emerson R, Aragon D, Hancock EB, et al. Estimating Influenza Disease Burden from Population-Based Surveillance Data in the United States. PLOS ONE. 2015 Mar 4;10(3):e0118369. doi:10.1371/journal.pone.0118369
